# Empagliflozin in patients admitted to hospital with COVID-19 (RECOVERY): a randomised, controlled, open-label, platform trial

**DOI:** 10.1101/2023.04.13.23288469

**Authors:** Peter W Horby, Natalie Staplin, Leon Peto, Jonathan R Emberson, Mark Campbell, Guilherme Pessoa-Amorim, Buddha Basnyat, Louise Thwaites, Rogier Van Doorn, Raph L Hamers, Jeremy Nel, John Amuasi, Manisha Rawal, Dipansu Ghosh, Jonathan Douse, Fergus Hamilton, Anthony Kerry, Pinky Thu-Ta, John Widdrington, Chris Green, Purav Desai, Richard Stewart, Nguyen Thanh Phong, J Kenneth Baillie, Maya H Buch, Saul N Faust, Thomas Jaki, Katie Jeffery, Edmund Juszczak, Marian Knight, Wei Shen Lim, Alan Montgomery, Aparna Mukherjee, Andrew Mumford, Kathryn Rowan, Guy Thwaites, Marion Mafham, Richard Haynes, Martin J Landray, RECOVERY Collaborative Group

**Author notes:** Correspondence to: Prof Peter W Horby and Prof Martin J Landray, RECOVERY Central Coordinating Office, Richard Doll Building, Old Road Campus, Roosevelt Drive, Oxford OX3 7LF, United Kingdom. The writing committee and trial steering committee are listed at the end of this manuscript and a complete list of collaborators in the Randomised Evaluation of COVID-19 Therapy (RECOVERY) trial is provided in the Supplementary Appendix.

## Abstract

**Background:** Empagliflozin has been proposed as a treatment for COVID-19 on the basis of its anti-inflammatory, metabolic and haemodynamic effects.

**Methods:** In this randomised, controlled, open-label trial, several possible treatments are compared with usual care in patients hospitalised with COVID-19. Eligible and consenting adults were randomly allocated in a 1:1 ratio to either usual standard of care alone or usual standard of care plus empagliflozin 10mg once daily for 28 days or until discharge using web-based simple (unstratified) randomisation with allocation concealment. The primary outcome was 28-day mortality. On 3 March the independent data monitoring committee recommended that the investigators review the data and recruitment was consequently stopped on 7 March. The trial is registered with ISRCTN (50189673) and clinicaltrials.gov (NCT04381936).

**Findings:** Between 8 July 2021 and 6 March 2023, 4271 patients were randomly allocated to receive either empagliflozin (2113 patients) or usual care alone (2158 patients). Overall, 289 (14%) patients allocated to empagliflozin and 307 (14%) patients allocated to usual care died within 28 days (rate ratio 0.96; 95% confidence interval [CI] 0.82-1.13; p=0.64). There was no evidence of significant differences in duration of hospitalisation (median 8 days vs. 8 days) or the proportion of patients discharged from hospital alive within 28 days (79% vs. 78%; rate ratio 1.03; 95% CI 0.96-1.10; p=0.44). Among those not on invasive mechanical ventilation at baseline, there was no evidence of a significant difference in the proportion meeting the composite endpoint of invasive mechanical ventilation or death (16% vs. 17%; risk ratio 0.95; 95% CI 0.84-1.08; p=0.44).

**Interpretation:** In adults hospitalised with COVID-19, empagliflozin was not associated with reductions in 28-day mortality, duration of hospital stay, or risk of progressing to invasive mechanical ventilation or death.

**Funding:** UK Research and Innovation (Medical Research Council) and National Institute of Health Research (Grant ref: MC_PC_19056), and Wellcome Trust (Grant Ref: 222406/Z/20/Z).

**Trial registration:** ClinicalTrials.gov NCT04381936 https://clinicaltrials.gov/ct2/show/NCT04381936

ISRCTN50189673 http://www.isrctn.com/ISRCTN50189673

## INTRODUCTION

Patients with cardiometabolic diseases (such as heart failure, diabetes and chronic kidney disease) are at increased risk of hospitalization and death from COVID-19. Sodium glucose co-transporter 2 inhibitors (SGLT2i) have been shown to reduce cardiovascular and kidney events in patients with cardiometabolic diseases.^1^ The precise mechanisms of such benefit are not known, but SGLT2i appear to favourably modify some pathways that are dysregulated in acute illnesses like COVID-19.

Inflammation is a key feature of severe COVID-19. Markedly raised levels of inflammatory markers such as C-reactive protein, ferritin, interleukin-6 (IL-6) and other cytokines are observed in severe cases and are associated with poor outcomes.^2,3^ Corticosteroids, IL-6 inhibitors and Janus kinase (JAK) inhibitors have been shown to reduce mortality in patients with severe COVID-19.^4-6^ Together these results show that inflammation is modifiable and anti-inflammatory therapy can improve clinical outcomes. SGLT2i can reduce inflammation,^7,8^ including via attenuation of the nucleotide binding domain (NOD)-like pyrin domain 3 (NLRP3) inflammasome, which correlates with disease severity in COVID-19.^9,10^ A meta-analysis of 26 trials in patients with type 2 diabetes mellitus showed a reduced risk of pneumonia and septic shock among patients allocated SGLT2i.^11^ In addition, SGLT2i inhibit glycolysis and stimulate lipolysis which may create a less favourable energetic environment for viruses like COVID-19,^12-14^ and improve endothelial function.^15^

The DARE-19 trial compared dapagliflozin 10mg once daily with placebo in 1250 patients hospitalised with COVID-19 who had at least one cardiometabolic risk factor but were not critically ill.^16^ The primary outcome of new or worsened organ dysfunction or death occurred in 70 (11%) of the dapaglifozin group versus 86 (14%) in the placebo group (hazard ratio [HR] 0.80, 95% CI 0.58-1.10). The dual primary outcome of improvement in clinical status by day 30 was also not significantly affected (win ratio 1.09, 95% CI 0.97-1.22). Dapagliflozin was well-tolerated and appeared safe (with fewer serious adverse events reported in the dapaglifozin group compared to placebo). Here we report the results of a large randomised controlled trial of empagliflozin in patients hospitalised with COVID-19.

## METHODS

### Study design and participants

The Randomised Evaluation of COVID-19 therapy (RECOVERY) trial is an investigator-initiated, individually randomised, controlled, open-label, adaptive platform trial to evaluate the effects of potential treatments in patients hospitalised with COVID-19. Details of the trial design and results for other possible treatments (dexamethasone, hydroxychloroquine, lopinavir-ritonavir, azithromycin, tocilizumab, convalescent plasma, colchicine, aspirin, casirivimab plus imdevimab, baricitinib, and high-dose corticosteroids in hypoxic patients not requiring ventilatory support) have been published previously.^6,17-26^ The trial is underway at hospital organisations in the United Kingdom supported by the National Institute for Health and Care Research Clinical Research Network, as well as in South and Southeast Asia and Africa. Of these, 118 hospitals in the UK, 5 in Nepal, 4 in Indonesia, 2 in Vietnam, 4 in South Africa, 1 in Ghana and 5 in India enrolled participants in the evaluation of empagliflozin (appendix pp 2-31). The trial is coordinated by the Nuffield Department of Population Health at University of Oxford (Oxford, UK), the trial sponsor. The trial is conducted in accordance with the principles of the International Conference on Harmonisation–Good Clinical Practice guidelines and approved by the UK Medicines and Healthcare products Regulatory Agency (MHRA) and the Cambridge East Research Ethics Committee (ref: 20/EE/0101). The protocol, statistical analysis plan, and additional information are available on the study website www.recoverytrial.net.

Patients admitted to hospital were eligible for the study if they had clinically suspected or laboratory confirmed SARS-CoV-2 infection and no medical history that might, in the opinion of the attending clinician, put the patient at significant risk if they were to participate in the trial. Children (age <18 years) and pregnant women were not eligible for randomisation due to limited data on use of empagliflozin in these groups such that it was not possible to make an evidence-based benefit-risk assessment. Patients with type 1 diabetes mellitus (or post-pancreatectomy diabetes), a history of ketoacidosis or type 2 diabetes mellitus with ketosis at the time of recruitment were ineligible for the comparison of empagliflozin vs. usual care (further details in appendix pp 32-33). Written informed consent was obtained from all patients, or a legal representative if patients were too unwell or unable to provide consent.

### Randomisation and masking

Baseline data were collected using a web-based case report form that included demographics, level of respiratory support, major comorbidities, suitability of the study treatment for a particular patient, SARS-CoV-2 vaccination status, and treatment availability at the study site (appendix pp 43-45). For some patients, empagliflozin was unavailable at the hospital at the time of enrolment or was considered by the managing physician to be either definitely indicated or definitely contraindicated. These patients were excluded from the randomised comparison between empagliflozin and usual care. Eligible and consenting adult patients were assigned in a 1:1 ratio to either usual standard of care or usual standard of care plus empagliflozin using web-based simple (unstratified) randomisation with allocation concealed until after randomisation (appendix pp 41-43). Patients allocated to empagliflozin were to receive 10mg orally daily for 28 days in total or until discharge, whichever occurred earlier. Investigators were instructed how to identify and manage ketosis that may develop during treatment with empagliflozin (appendix p 32-33).

As a platform trial, and in a factorial design, patients could be simultaneously randomised to other treatment groups: i) baricitinib versus usual care, ii) higher-dose corticosteroids versus usual care, iii) sotrovimab versus usual care, iv) molnupiravir versus usual care, and v) nirmatrelvir-ritonavir versus usual care (appendix pp 41-42). Participants and local study staff were not masked to the allocated treatment. Other than members of the Data Monitoring Committee, all individuals involved in the trial were masked to aggregated outcome data while recruitment and 28-day follow-up were ongoing.

### Procedures

A single online follow-up form was completed when participants were discharged, had died or at 28 days after randomisation, whichever occurred earliest (appendix pp 47-55). Information was recorded on adherence to allocated study treatment, receipt of other COVID-19 treatments, duration of admission, receipt of respiratory or renal support, and vital status (including cause of death). In addition, in the UK, routine healthcare and registry data were obtained including information on vital status (with date and cause of death), discharge from hospital, receipt of respiratory support, or renal replacement therapy. For sites outside the UK a further case report form (appendix pp 56-57) collected vital status at day 28 (if not already reported on follow-up form).

### Outcomes

Outcomes were assessed at 28 days after randomisation, with further analyses specified at 6 months. The primary outcome was all-cause mortality at 28 days. Secondary outcomes were time to discharge from hospital, and, among patients not on invasive mechanical ventilation at randomisation, invasive mechanical ventilation (including extra-corporal membrane oxygenation) or death. Prespecified subsidiary clinical outcomes were use of non-invasive respiratory support, time to successful cessation of invasive mechanical ventilation (defined as cessation of invasive mechanical ventilation within, and survival to, 28 days), use of renal dialysis or haemofiltration, cause-specific mortality, bleeding events, thrombotic events, major cardiac arrhythmias, thrombotic and bleeding events, other infections and metabolic complications (including ketoacidosis). Information on suspected serious adverse reactions was collected in an expedited fashion to comply with regulatory requirements.

### Sample size and role of the independent Data Monitoring Committee

The intention for this comparison was to continue recruitment until sufficient primary outcomes had accrued to have 90% power to detect a proportional risk reduction of 20% at a two-sided significance level of 0.01.

The independent Data Monitoring Committee reviewed unblinded analyses of the study data and any other information considered relevant to the trial at intervals of around 2-3 months (depending on speed of enrolment) and was charged with determining if, in their view, the randomised comparisons in the study provided evidence on mortality that was strong enough (with a range of uncertainty around the results that was narrow enough) to affect national and global treatment strategies (appendix p 58).

On 3 March 2023, the Data Monitoring Committee recommended that the investigators review the unblinded data from the empagliflozin comparison (appendix p 59). Consequently, recruitment to the empagliflozin comparison was closed on 7 March 2023.

### Statistical Analysis

The primary analysis for all outcomes was by intention-to-treat comparing patients randomised to empagliflozin with patients randomised to usual care but for whom empagliflozin was both available and suitable as a treatment. For the primary outcome of 28-day mortality, the hazard ratio from an age- and respiratory status-adjusted Cox model was used to estimate the mortality rate ratio. We constructed Kaplan-Meier survival curves to display cumulative mortality over the 28-day period. We used the same Cox regression method to analyse time to hospital discharge and successful cessation of invasive mechanical ventilation, with patients who died in hospital right-censored on day 29. Median time to discharge was derived from Kaplan-Meier estimates. For the pre-specified composite secondary outcome of progression to invasive mechanical ventilation or death within 28 days (among those not receiving invasive mechanical ventilation at randomisation), and the subsidiary clinical outcomes of receipt of ventilation and use of haemodialysis or haemofiltration, the precise dates were not available and so a log-binomial regression model was used to estimate the risk ratio adjusted for age and respiratory status.

Prespecified subgroup analyses were performed for the primary outcome using the statistical test of interaction (test for heterogeneity or trend), in accordance with the prespecified analysis plan, defined by the following characteristics at randomisation: age, sex, ethnicity, level of respiratory support, days since symptom onset, and use of corticosteroids (appendix p 139).

Estimates of rate and risk ratios are shown with 95% confidence intervals. All p-values are 2-sided and are shown without adjustment for multiple testing. The full database is held by the study team which collected the data from study sites and performed the analyses at the Nuffield Department of Population Health, University of Oxford (Oxford, UK).

Analyses were performed using SAS version 9.4 and R version 3.4. The trial is registered with ISRCTN (50189673) and clinicaltrials.gov (NCT04381936).

## RESULTS

Between 8 July 2021 and 6 March 2023, 4271 (74%) of 5740 patients enrolled into the RECOVERY trial were eligible to be randomly allocated to empagliflozin (i.e. empagliflozin was available in the hospital at the time and the attending clinician was of the opinion that the patient had no known indication for or contraindication to empagliflozin, figure 1). The characteristics of the 1469 patients enrolled into the RECOVERY trial during this period but not included in the empagliflozin comparison are shown in webtable 1. 2113 patients were randomly allocated to empagliflozin and 2158 were randomly allocated to usual care. The mean age of study participants in this comparison was 61.5 years (SD 16.4) and the median time since symptom onset was 8 days (IQR 5 to 11 days). At randomisation, 3842 (90%) patients were receiving corticosteroids, about one-quarter were receiving remdesivir and about one-quarter had received tocilizumab (table 1).

**Table 1:** Baseline characteristics

|  | Treatment allocation |  |
| --- | --- | --- |
|  | Empagliflozin<br>(n=2113) | Usual care<br>(n=2158) |
| Age, years | 61.1 (16.3) | 61.8 (16.4) |
| <70 | 1412 (67%) | 1393 (65%) |
| ≥70 to <80 | 434 (21%) | 479 (22%) |
| ≥80 | 267 (13%) | 286 (13%) |
| Sex |  |  |
| Male | 1326 (63%) | 1339 (62%) |
| Female* | 787 (37%) | 819 (38%) |
| Country |  |  |
| Ghana | 2 (<0.5%) | 2 (<0.5%) |
| India | 24 (1%) | 19 (1%) |
| Indonesia | 68 (3%) | 68 (3%) |
| Nepal | 139 (7%) | 119 (6%) |
| South Africa | 10 (<0.5%) | 14 (1%) |
| Vietnam | 40 (2%) | 53 (2%) |
| UK | 1830 (87%) | 1883 (87%) |
| Race |  |  |
| White | 1557 (74%) | 1607 (74%) |
| Black, Asian, and minority ethnic | 361 (17%) | 330 (15%) |
| Unknown | 195 (9%) | 221 (10%) |
| Number of days since symptom onset | 8 (5-11) | 8 (5-12) |
| Number of days since hospitalisation | 2 (1-3) | 2 (1-3) |
| Respiratory support received |  |  |
| None | 255 (12%) | 260 (12%) |
| Simple oxygen | 1317 (62%) | 1383 (64%) |
| Non invasive ventilation | 512 (24%) | 500 (23%) |
| Invasive mechanical ventilation | 29 (1%) | 15 (1%) |
| Biochemistry |  |  |
| C-reactive protein, mg/L | 83 (39-148) | 85 (38-151) |
| Creatinine, umol/L | 75 (62-94) | 78 (63-96) |
| Previous diseases |  |  |
| Diabetes | 333 (16%) | 356 (16%) |
| Heart disease | 471 (22%) | 455 (21%) |
| Chronic lung disease | 533 (25%) | 508 (24%) |
| Tuberculosis | 9 (<0.5%) | 7 (<0.5%) |
| HIV | 21 (1%) | 13 (1%) |
| Severe liver disease† | 20 (1%) | 21 (1%) |
| Severe kidney impairment‡ | 66 (3%) | 80 (4%) |
| Any of the above | 1014 (48%) | 1039 (48%) |
| SARS-CoV-2 PCR test result |  |  |
| Positive | 2046 (97%) | 2097 (97%) |
| Negative | 12 (1%) | 10 (<0.5%) |
| Unknown | 55 (3%) | 51 (2%) |
| Received a COVID-19 vaccine | 1412 (67%) | 1453 (67%) |
| Use of other treatments |  |  |
| Corticosteroids | 1910 (90%) | 1932 (90%) |
| Remdesivir | 541 (26%) | 547 (25%) |
| Tocilizumab | 504 (24%) | 491 (23%) |
| Plan to use tocilizumab within the next 24 hours | 208 (10%) | 240 (11%) |

Empagliflozin for COVID-19
|  | Treatment allocation |  |
| --- | --- | --- |
|  | Empagliflozin<br>(n=2113) | Usual care<br>(n=2158) |
| Other randomly assigned treatments |  |  |
| Baricitinib | 554 (26%) | 591 (27%) |
| High dose steroids | 160 (8%) | 161 (7%) |
| Sotrovimab | 185 (9%) | 194 (9%) |
| Molnupiravir | 143 (7%) | 133 (6%) |
| Nirmatrelvir-ritonavir | 19 (1%) | 28 (1%) |

Results are count (%), mean ± standard deviation, or median (inter-quartile range). \*Includes 0 pregnant women.
†Defined as requiring ongoing specialist care. ‡Defined as estimated glomerular filtration rate $<30$ mL/min/1.73m<sup>2</sup>

**Figure 1:**
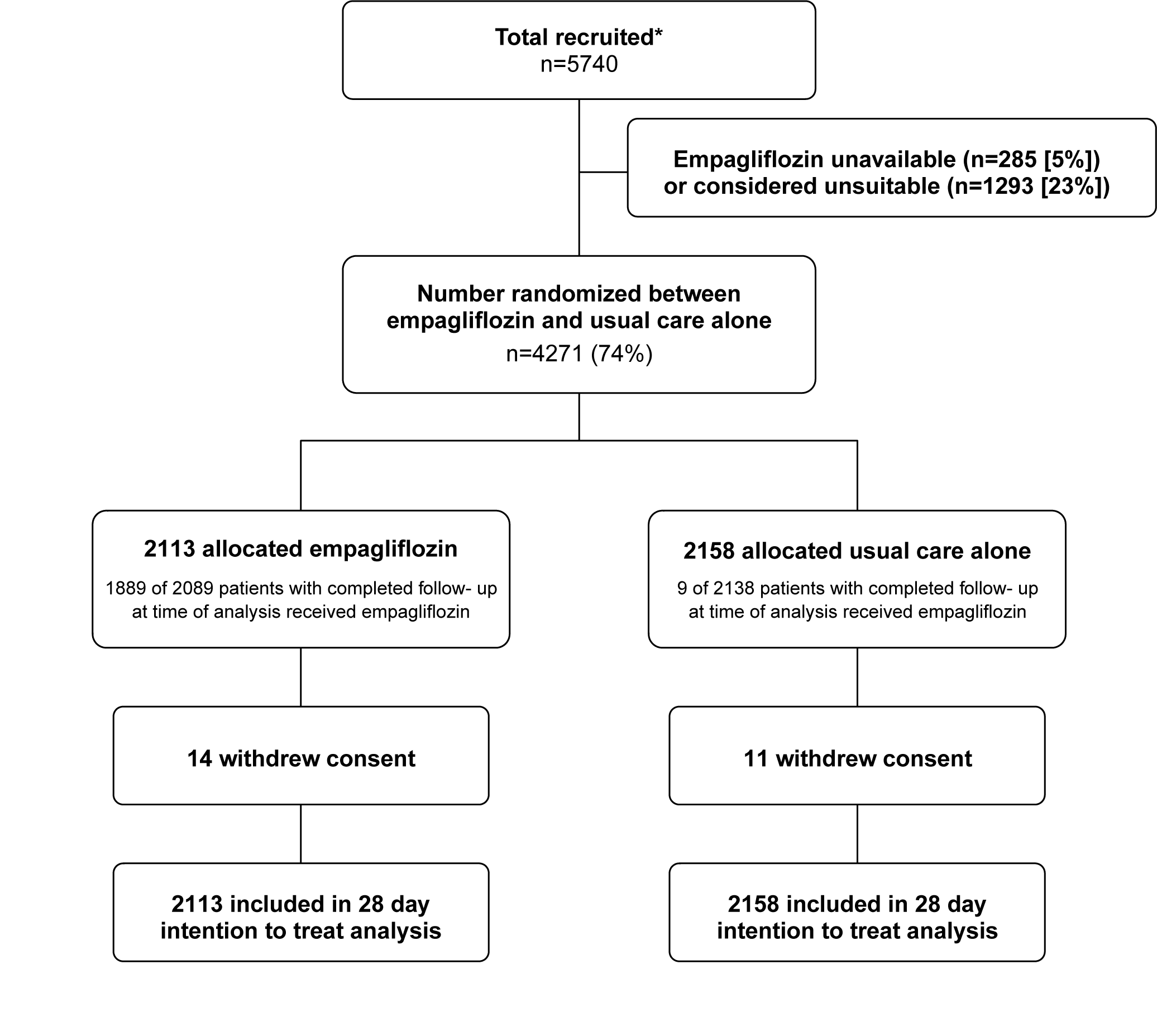
Trial profile. ITT=intention to treat. * Number recruited overall during period that adult participants could be recruited into empagliflozin comparison. Of the 4271 randomised to empagliflozin vs usual care, 1145 were additionally randomised to baricitinib vs usual care (554 [26%] of the empagliflozin group vs 591 [27%] of the usual care group); 321 were additionally randomised to higher-dose corticosteroids vs usual care (160 [8%] of the empagliflozin group vs 161 [7%] of the usual care group); 379 were additionally randomised to sotrovimab vs usual care (185 [9%] of the empagliflozin group vs 194 [9%] of the usual care group); 276 were additionally randomised to molnupiravir vs usual care (143 [7%] of the empagliflozin group vs 133 [6%] of the usual care group); and 47 patients were additionally randomised to nirmatrelvir-ritonavir vs usual care (19 [1%] of the empagliflozin group vs 28 [1%] of the usual care group).

The follow-up form was completed for 2089 (99%) patients in the empagliflozin group and 2138 (99%) patients in the usual care group. Among patients with a completed follow-up form, 1889 (90%) allocated to empagliflozin received at least one dose and, of these, 1321 (70%) received it on most (≥90%) days of their admission (or until 28 days after randomisation if not discharged sooner) (figure 1; webtable 2). By comparison, <1% of those allocated to usual care alone received any dose of empagliflozin. Use of other treatments for COVID-19 was similar among patients allocated empagliflozin and among those allocated usual care (webtable 2).

**Table 2:** Effect of allocation to empagliflozin on key study outcomes

|  | Treatment allocation |  | RR (95% CI) | p value |
| --- | --- | --- | --- | --- |
|  | Empagliflozin<br>(n=2113) | Usual care<br>(n=2158) |  |  |
| **Primary outcome:** |  |  |  |  |
| 28-day mortality | 289 (14%) | 307 (14%) | 0.96 (0.82-1.13) | 0.64 |
| **Secondary outcomes:** |  |  |  |  |
| Median time to being discharged alive, days | 8 (5 to 19) | 8 (5 to 20) |  |  |
| Discharged from hospital within 28 days | 1678 (79%) | 1677 (78%) | 1.03 (0.96-1.10) | 0.44 |
| Receipt of invasive mechanical ventilation or death* | 338/2084 (16%) | 371/2143 (17%) | 0.95 (0.84-1.08) | 0.44 |
| Invasive mechanical ventilation | 130/2084 (6%) | 133/2143 (6%) | 0.97 (0.77-1.21) | 0.77 |
| Death | 274/2084 (13%) | 302/2143 (14%) | 0.96 (0.84-1.11) | 0.59 |
| **Subsidiary clinical outcomes** |  |  |  |  |
| Receipt of ventilation† | 245/1567 (16%) | 259/1641 (16%) | 1.00 (0.85-1.17) | 0.97 |
| Non-invasive ventilation | 237/1572 (15%) | 252/1643 (15%) | 0.99 (0.84-1.16) | 0.90 |
| Invasive mechanical ventilation | 51/1567 (3%) | 49/1641 (3%) | 1.09 (0.74-1.60) | 0.66 |
| Successful cessation of invasive mechanical ventilation‡ | 10/29 (35%) | 6/15 (40%) | 0.67 (0.24-1.85) | 0.44 |
| Renal replacement therapy§ | 45/2103 (2%) | 44/2146 (2%) | 0.96 (0.64-1.45) | 0.86 |

RR=Rate Ratio for the outcomes of 28-day mortality and hospital discharge, and risk ratio for the outcome of receipt of invasive mechanical ventilation or death (and its subcomponents). CI=confidence interval. \*Analyses exclude those on invasive mechanical ventilation at randomization. †Analyses exclude those on any form of ventilation at randomisation. ‡Analyses restricted to those on invasive mechanical ventilation at randomisation. §Analyses exclude those on haemodialysis or haemofiltration at randomisation.

Primary and secondary outcome data are known for >99% of randomly assigned patients. There was no evidence of a significant difference in the proportion of patients who met the primary outcome of 28-day mortality between the two randomised groups (289 [14%] patients in the empagliflozin group vs. 307 [14%] patients in the usual care group; hazard ratio 0.96; 95% confidence interval [CI], 0.82-1.13; p=0.64; figure 2). We observed similar results across all pre-specified sub-groups (figure 3), except among the small group of patients not requiring oxygen at baseline or not receiving corticosteroids among whom there were very few (about 20) events.

**Figure 2:**
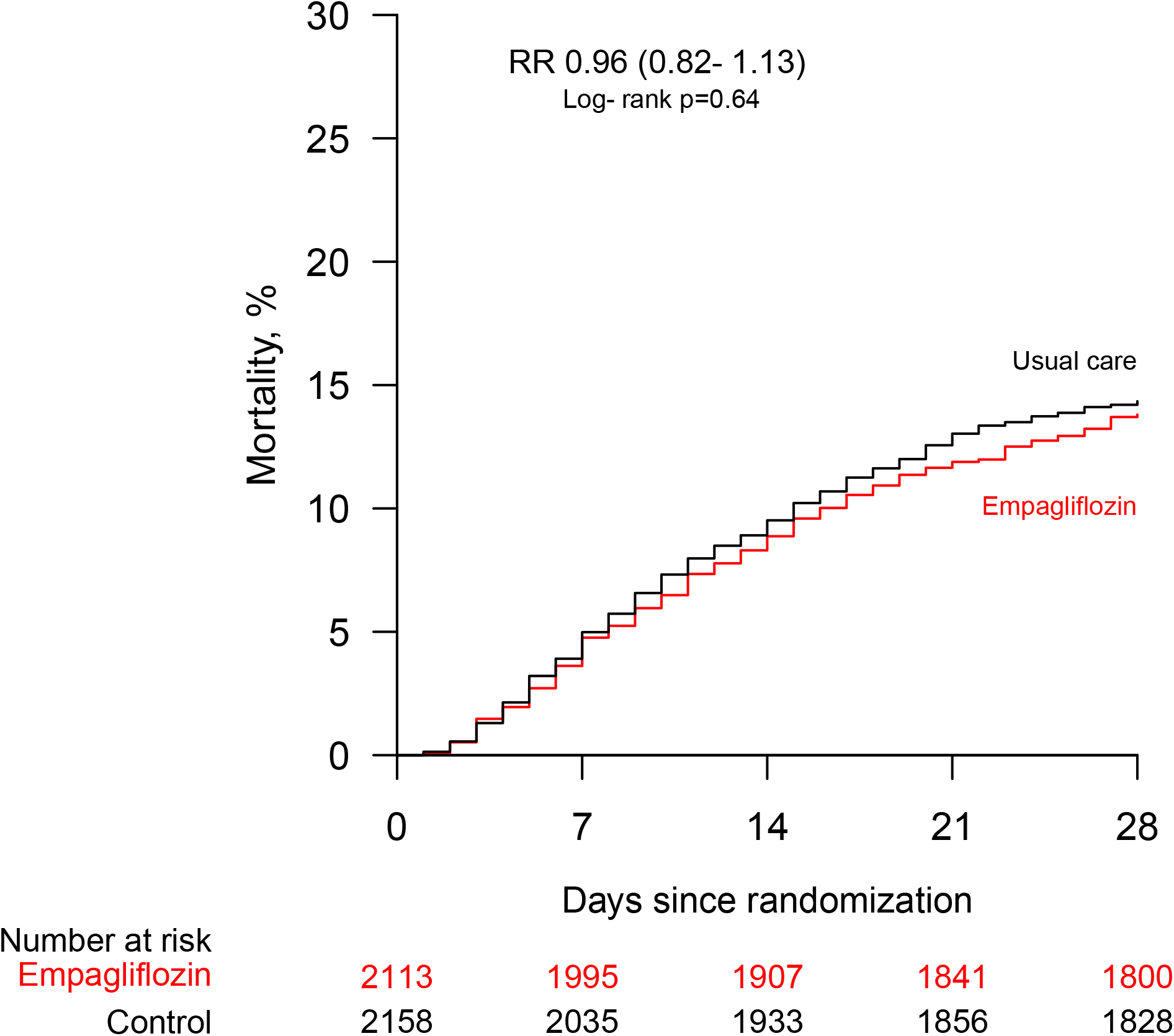
Effect of allocation to empagliflozin on 28-day mortality.

**Figure 3:**
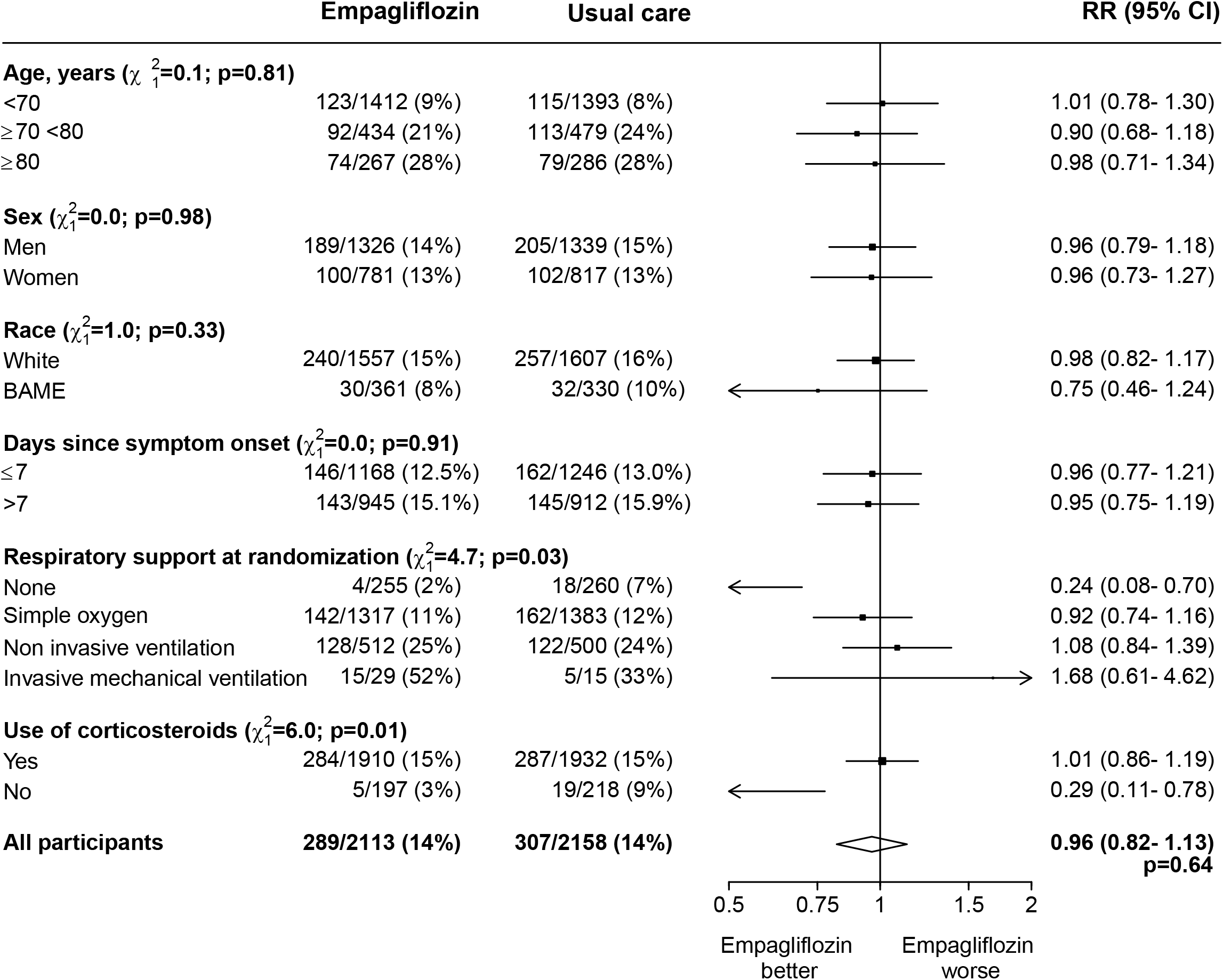
Effect of allocation to empagliflozin on 28-day mortality by baseline characteristics. Subgroup−specific rate ratio estimates are represented by squares (with areas of the squares proportional to the amount of statistical information) and the lines through them correspond to the 95% CIs. The ethnicity, days since onset and use of corticosteroids subgroups exclude those with missing data, but these patients are included in the overall summary diamond.

The median time to discharge from hospital alive was 8 days in both groups and there was no evidence of a significant difference in the probability of being discharged alive within 28 days (79 vs. 78%, rate ratio 1.03, 95% CI 0.96 to 1.10, p=0.44) (table 2). Among those not on invasive mechanical ventilation at baseline, the number of patients progressing to the pre-specified composite secondary outcome of invasive mechanical ventilation or death was similar in both groups (16% vs. 17%, risk ratio 0.95, 95% CI 0.84 to 1.08, p=0.44). Similar results were seen in all pre-specified subgroups of patients (webfigure 1, webfigure 2).

We found no evidence of significant differences in the prespecified subsidiary clinical outcomes of cause-specific mortality (webtable 3), use of ventilation or successful cessation of invasive mechanical ventilation (table 2). We found no evidence of a significant difference in the incidence of acute kidney injury (defined as an increase in the pre-randomisation creatinine concentration of at least 50%), or need for renal dialysis or haemofiltration (table 2, webtable 4). The incidence of new cardiac arrhythmias, bleeding events, and non-coronavirus infections was also similar in the two groups (webtable 4). There were fewer thrombotic events among patients allocated empagliflozin compared to placebo (2.5% vs. 3.9%, absolute difference -1.4% [-0.4 to -2.5]; webtable 4). The incidence of metabolic complications were similar in the two groups, with reported ketoacidosis in 5 vs. 2 patients. There were two reports of a serious adverse reaction believed to be related to empagliflozin: both were ketosis without acidosis (including one patient without diabetes) and resolved rapidly on cessation of the drug.

## DISCUSSION

In this large, randomised trial involving over 4000 patients from 7 countries and nearly 600 deaths, allocation to empagliflozin was not associated with reductions in mortality, duration of hospitalisation, or the risk of being ventilated or dying for those not on ventilation at baseline. These results were consistent across prespecified subgroups of age, sex, race and duration of symptoms prior to randomisation.

The benefits of immunomodulatory therapies in patients with moderate to severe COVID-19 demonstrates the importance of inflammation in this patient group and empagliflozin was proposed as a treatment for COVID-19 partly based on its anti-inflammatory activity as well as purported benefits on endothelial function and cellular energy metabolism. The lack of evidence of benefit from empagliflozin in this large well-powered trial suggests that these properties of empagliflozin are either insufficient to produce a meaningful reduction in mortality risk or are not affecting the relevant pathways in moderate to severe COVID-19. There is weak evidence that there may be some benefit on 28-day mortality in patients not receiving a corticosteroid or not requiring oxygen, which are largely the same patients. However, this observation is based on a very small number of events, marginally significant tests for heterogeneity or trend, and is not supported by either of the secondary outcomes.

Only one other trial of an SGLT2i in COVID-19 has been reported to date. The DARE-19 trial recruited 1250 patients hospitalised (but not critically ill) with COVID-19 and with at least one cardiometabolic risk factor (i.e. hypertension, type 2 diabetes mellitus, atherosclerotic cardiovascular disease, heart failure or chronic kidney disease).^16^ There was no significant effect of dapagliflozin on either of the two dual primary outcomes (new or worsened organ function or death, and change in clinical status by day 30), although there were numerically fewer poor outcomes in dapagliflozin group, including 41 deaths compared to 54 in the placebo group. However, DARE-19 was not large enough to detect plausibly moderate benefits of treatment. Meta-analysing the results on mortality from DARE-19 and RECOVERY yields a summary relative risk of 0.93 (0.80-1.08). Trials of SGLT2i in chronic disease setting have found consistent evidence of a reduction in acute kidney injury (AKI): a meta-analysis of 13 large placebo-controlled trials including over 2000 AKI events reported a reduction of nearly one-quarter (relative risk 0.77, 95% CI 0.70-0.84) in this outcome.^1^ The RECOVERY trial did not find evidence of benefit (or harm) of empagliflozin on the risk of developing AKI in the acute setting where AKI may have already begun prior to randomisation.

Strengths of this trial included that it was randomised, had a large sample size, broad eligibility criteria, was international and more than 99% of patients were followed up for the primary outcome. However, detailed information on laboratory markers of inflammation and immune response was not collected, nor was information on radiological or physiological outcomes. Although this randomised trial is open label (i.e., participants and local hospital staff are aware of the assigned treatment), the outcomes are unambiguous and were ascertained without bias through linkage to routine health records in the large majority of patients.

The RECOVERY trial only studied patients who had been hospitalised with COVID-19 and, therefore, is not able to provide any evidence on the safety and efficacy of empagliflozin used in other patient groups. Due to the recommendation that empagliflozin be taken orally (and not via a gastric feeding tube), there were few patients recruited requiring invasive mechanical ventilation. Nevertheless, the reassuring safety findings in RECOVERY suggest that SGLT2i can be safely used in the acute setting and do not need to be routinely discontinued if there is an appropriate indication. These results show that the key risk of ketoacidosis can be safely mitigated with simple monitoring and advice to managing physicians.

In summary, the results of this large, randomised trial do not support the use of empagliflozin as a treatment for adults hospitalised with COVID-19.

## Data Availability

The protocol, consent form, statistical analysis plan, definition & derivation of clinical characteristics & outcomes, training materials, regulatory documents, and other relevant study materials are available online at www.recoverytrial.net. As described in the protocol, the Trial Steering Committee will facilitate the use of the study data and approval will not be unreasonably withheld. Deidentified participant data will be made available to bona fide researchers registered with an appropriate institution within 3 months of publication. However, the Steering Committee will need to be satisfied that any proposed publication is of high quality, honours the commitments made to the study participants in the consent documentation and ethical approvals, and is compliant with relevant legal and regulatory requirements (e.g. relating to data protection and privacy). The Steering Committee will have the right to review and comment on any draft manuscripts prior to publication. Data will be made available in line with the policy and procedures described at: https://www.ndph.ox.ac.uk/data-access. Those wishing to request access should complete the form at https://www.ndph.ox.ac.uk/files/about/data_access_enquiry_form_13_6_2019.docx and e-mailed to

## Contributors

This manuscript was initially drafted by the RH, PWH and MJL, further developed by the Writing Committee, and approved by all members of the trial steering committee. PWH and MJL vouch for the data and analyses, and for the fidelity of this report to the study protocol and data analysis plan. PWH, JKB, MB, MK, SNF, TJ, EJ, KJ, WSL, AMo, AMuk, AMum, JN, KR, GT, MM, RH, and MJL designed the trial and study protocol. MM, MC, G P-A, LP, RKJ, DG, JD, FH, AK, PT-T, JW, CG, PD, RS, the Data Linkage team at the RECOVERY Coordinating Centre, and the Health Records and Local Clinical Centre staff listed in the appendix collected the data. NS and JRE did the statistical analysis. All authors contributed to data interpretation and critical review and revision of the manuscript. PWH and MJL had access to the study data and had final responsibility for the decision to submit for publication.

## Writing Committee (on behalf of the RECOVERY Collaborative Group)

Peter W Horby, Natalie Staplin, Leon Peto, Jonathan R Emberson, Mark Campbell, Guilherme Pessoa-Amorim, Buddha Basnyat, Louise Thwaites, Rogier van Doorn, Raph L Hamers, Jeremy Nel, John Amuasi, Roshan Kumar Jha, Dipansu Ghosh, Jonathan Douse, Fergus Hamilton, Anthony Kerry, Pinky Thu-Ta, John Widdrington, Christopher A Green, Purav Desai, Richard Stewart, Nguyen Thanh Phong, J Kenneth Baillie, Maya Buch, Saul N Faust, Thomas Jaki, Edmund Juszczak, Katie Jeffery, Marian Knight, Wei Shen Lim, Alan Montgomery, Aparna Mukherjee, Andrew Mumford, Kathryn Rowan, Guy Thwaites, Marion Mafham, Richard Haynes, Martin J Landray.

## Data Monitoring Committee

Peter Sandercock, Janet Darbyshire, David DeMets, Robert Fowler, David Lalloo, Mohammed Munavvar, Janet Wittes.

## Acknowledgements

Above all, we would like to thank the thousands of patients who participated in this trial. We would also like to thank the many doctors, nurses, pharmacists, other allied health professionals, and research administrators at NHS hospital organisations across the whole of the UK, supported by staff at the National Institute of Health Research (NIHR) Clinical Research Network, NHS DigiTrials, Public Health England, Department of Health & Social Care, the Intensive Care National Audit & Research Centre, Public Health Scotland, National Records Service of Scotland, the Secure Anonymised Information Linkage (SAIL) at University of Swansea, and the NHS in England, Scotland, Wales and Northern Ireland.

The RECOVERY trial is supported by grants to the University of Oxford from UK Research and Innovation (UKRI) and NIHR (MC_PC_19056), the Wellcome Trust (Grant Ref: 222406/Z/20/Z) through the COVID-19 Therapeutics Accelerator, and by core funding provided by the NIHR Oxford Biomedical Research Centre, the Wellcome Trust, the Bill and Melinda Gates Foundation, the Foreign, Commonwealth and Development Office, Health Data Research UK, the Medical Research Council Population Health Research Unit, the NIHR Health Protection Unit in Emerging and Zoonotic Infections, and NIHR Clinical Trials Unit Support Funding. TJ is supported by a grant from UK Medical Research Council (MC_UU_0002/14). WSL is supported by core funding provided by NIHR Nottingham Biomedical Research Centre. Boehringer-Ingelheim supplied empagliflozin free of charge for use in this trial in countries outside the UK. Boehringer Ingelheim was given the opportunity to review the manuscript for medical and scientific consistency as it relates to Boehringer Ingelheim substances, as well as intellectual property considerations.

The views expressed in this publication are those of the authors and not necessarily those of the NHS, the NIHR, or the UK Department of Health and Social Care.

